# Design and Analysis of Outcomes following SARS-CoV-2 Infection in Veterans

**DOI:** 10.1101/2022.08.23.22279120

**Authors:** Valerie A. Smith, Theodore S. Z. Berkowitz, Paul Hebert, Edwin S. Wong, Meike Niederhausen, John A. Pura, Kristin Berry, Pamela Green, Anna Korpak, Alexandra Fox, Aaron Baraff, Alex Hickok, Troy A Shahoumian, Amy S.B. Bohnert, Denise Hynes, Edward J. Boyko, George N. Ioannou, Theodore J. Iwashyna, C. Barrett Bowling, Ann M. O’Hare, Matthew L. Maciejewski

**Author notes:** <u>Corresponding Author:</u> Matthew L. Maciejewski, Ph.D., Department of Population Health Sciences, Duke University Medical Center, Durham, NC 27705.

## Abstract

**Background:** Understanding how SARS-CoV-2 infection impacts long-term patient outcomes requires identification of comparable persons with and without infection. We report the design and implementation of a matching strategy employed by the Department of Veterans Affairs’ (VA) COVID-19 Observational Research Collaboratory (CORC) to develop comparable cohorts of SARS-CoV-2 infected and uninfected persons for the purpose of inferring potential causative long-term adverse effects of SARS-CoV-2 infection in the Veteran population.

**Methods:** In a retrospective cohort study, we identified VA health care system patients who were and were not infected with SARS-CoV-2 on a rolling monthly basis. We generated matched cohorts utilizing a combination of exact and time-varying propensity score matching based on electronic health record (EHR)-derived covariates that can be confounders or risk factors across a range of outcomes.

**Results:** From an initial pool of 126,689,864 person-months of observation, we generated final matched cohorts of 208,536 Veterans infected between March 2020-April 2021 and 3,014,091 uninfected Veterans. Matched cohorts were well-balanced on all 38 covariates used in matching after excluding patients for: no VA health care utilization; implausible age, weight, or height; living outside of the 50 states or Washington, D.C.; prior SARS-CoV-2 diagnosis per Medicare claims; or lack of a suitable match. Most Veterans in the matched cohort were male (88.3%), non-Hispanic (87.1%), white (67.2%), and living in urban areas (71.5%), with a mean age of 60.6, BMI of 31.3, Gagne comorbidity score of 1.4 and a mean of 2.3 CDC high-risk conditions. The most common diagnoses were hypertension (61.4%), diabetes (34.3%), major depression (32.2%), coronary heart disease (28.5%), PTSD (25.5%), anxiety (22.5%), and chronic kidney disease (22.5%).

**Conclusions:** This successful creation of matched SARS-CoV-2 infected and uninfected patient cohorts from the largest integrated health system in the United States will support cohort studies of outcomes derived from EHRs and sample selection for qualitative interviews and patient surveys. These studies will increase our understanding of the long-term outcomes of Veterans who were infected with SARS-CoV-2.

## Background

The U.S. faces continued infections with Severe Acute Respiratory Syndrome Coronavirus 2 (SARS-CoV-2) following earlier waves in 2020 and 2021. Numerous studies have examined short-term symptoms, hospitalization, and death(1, 2) among patients infected with SARS-CoV-2 both before and after vaccine availability. The coronavirus disease 2019 (COVID-19) pandemic caused unprecedented disruptions across a wide range of domains relevant to health and health care including to health systems and care processes, informal family support, long-term care, safety net programs, social organizations, and economic stability, making it challenging to isolate the direct impact of infection with SARS-CoV-2 on individual health. Thus, more work is needed to characterize the direct long-term health effects of SARS-CoV-2 infection distinct from these systemic disruptions.

To research the long-term health consequences of COVID-19, the U.S. Department of Veterans Affairs (VA) provided resources in May 2021 to create the COVID-19 Outcomes Research Collaboratory (CORC). The main purposes of this program are investigating long-term health outcomes associated with SARS-CoV-2 infection using national electronic health record (EHR) data and survey research to assess factors not well captured by the EHR. The focus of this paper is on the study design utilized to facilitate EHR research through cohort construction of Veterans with SARS-CoV-2 infection and well-matched controls, with matching for a wide range of covariates potentially associated with the exposure (SARS-CoV-2 infection) and outcomes. The creation of these populations will enable study of outcomes associated with this infection and potential mediating factors using observational research methods applied to both retrospective EHR and prospective survey-based data.

To best identify potential causal associations, we selected the target trial emulation approach to evaluate the causal effect of SARS-CoV-2 infection on long-term health-related outcomes. This approach is intended to minimize sources of bias, including observed and unobserved confounding and immortal time bias in estimating the effect of SARS-CoV-2 infection.(3, 4) To address unobserved confounding and selection bias, we matched Veterans infected with SARS-CoV-2 to similar contemporaneous uninfected Veterans before comparing a broad array of outcomes available in the VA’s comprehensive longitudinal EHR.

Analyses based on the EHR will be supplemented with a prospective longitudinal survey in which a subset of matched cohorts of infected and uninfected persons will be invited to participate. Longitudinal survey responses will provide more detailed information on patient-reported outcomes that will complement outcomes ascertained from the EHR. Selection and matching of survey samples will be conducted in such a way to ensure covariate balance is maintained in the survey subsample. In addition, source data from the EHR and survey will be used to guide purposive sampling for a prospective qualitative study to understand the diversity of experiences of Veterans infected with SARS-CoV-2.

Designing a matched cohort study to address a wide array of EHR-based outcomes and embedded survey subsets requires a more inclusive consideration of confounding than when estimating the effect of SARS-CoV-2 infection on a single outcome. This protocol paper describes the design and methodological approach to identify a matched cohort of comparable patients infected and not infected with SARS-CoV-2. This matched cohort will be used in future research to analyze the effect of SARS-CoV-2 infection on clinical, functional, and economic outcomes among Veterans. This work could also inform and support efforts by other groups interested in creating matched cohorts to address a wide range of unanswered questions related to SARS-CoV-2 infection.

## Methods

### Study Design and Data

We designed a retrospective cohort study of EHR-based outcomes with a non-equivalent comparator of uninfected Veterans. To facilitate measurement of patient-reported outcomes, this retrospective cohort is paired with an embedded smaller post-only survey-based prospective cohort study. In both components, comparator non-equivalence was reduced by generating matched cohorts.

As described previously,(5) we assembled a cohort of VA enrollees who tested positive for SARS-CoV-2 RNA in a respiratory specimen within the VA system based on polymerase chain reaction (PCR) tests as well as those with such tests performed outside the VA but documented in VA records as identified by the VA National Surveillance Tool between March 1, 2020 and April 30, 2021. The earliest date of a documented positive test was taken as each patient’s date of infection. We included only those Veterans who had an assigned VA primary care team (e.g., Patient Aligned Care Team) or at least one VA primary care clinic visit in the two-year period prior to infection to minimize missingness in EHR-based covariates that are generated from health system interaction. Cohorts were identified sequentially on a monthly basis, with assignment to a particular month for cases based on the date of the positive test. VA-enrolled Veterans without a positive test prior to or during the month who met the same inclusion criteria were considered uninfected potential comparators for that month. The uninfected control group members were eligible for repeated sampling and matching with replacement until they had a positive test. To avoid misclassification of first infection date based on a positive test, infected Veterans with COVID-19-related diagnostic codes (ICD-10: B97.29, U07.1, J12.82, Z86.16) listed in fee-for-service Medicare claims 15 or more days before their VA test were excluded. In addition, Veterans from the uninfected comparator group with any such diagnostic codes were excluded from sampling for matching in the month the COVID-19-related code arose and any months thereafter.

We developed 14 separate monthly patient cohorts—one for each month (March 2020-April 2021) —for the purpose of defining index dates and matching covariates. For example, the March 2020 cohort included all VA enrollees with an initial positive test during March 2020 and all VA enrollees who were alive as of March 1, 2020 and had not been infected prior to April 1, 2020. SARS-CoV-2-infected patients were included as potential comparator patients in months before infection. In a given month, uninfected Veterans could be matched to multiple infected Veterans in that same month and uninfected Veterans could be included in multiple month-specific cohorts as long as they remained uninfected and continued to meet other eligibility criteria. To minimize immortal time bias, the index date was defined as the date of the earliest positive test for SARS-CoV-2-infected Veterans and as the 1^st^ day of the relevant month for uninfected Veterans.(6) Each patient’s index date served as the anchor for defining matching covariates (with covariate construction starting 14 days prior to the positive test date for infected patients), based on EHR data from the prior two years.

### Matching Specification

Our goal was to conduct many-to-one matching that would maximize retention of infected patients for external validity and covariate balance for internal validity. A priori, we defined a suitable matching strategy as one that would result in <5% attrition of the infected cohort and achieve covariate balance among the selected covariates for matching based on standardized differences <0.1.(7)

Coarsened exact matching (CEM) was initially attempted. Covariates used for matching were derived iteratively at a single point in time (summer 2021) with the understanding that the evidence base about causes and consequences of COVID-19 was (and is) evolving rapidly. In collaboration with clinician-investigators (see left column, Appendix 1), we identified a broad list of demographic, clinical, and health care utilization measures hypothesized to be either risk factors for pre-specified outcomes alone (e.g., survival, depression, total VA costs, disability, financial toxicity) or confounders associated with both infection and outcomes.(8)

To minimize sample loss when attempting to match on many covariates in CEM,(9) the five physician principal investigators then worked together to prioritize covariates for the final matching specification (see right column, Appendix 1). Modified coarsened exact matching was then implemented using this prioritized set of covariates. However, a suitable exact match could not be identified for 53.7% of infected Veterans, so we reverted to a form of combined exact and calendar time-specific propensity score matching,(10) with cohorts identified by index month.

In a two-step process, infected patients were exact matched to uninfected controls based on index month, sex, immunosuppressive medication use (binary), state of residence, and COVID-19 vaccination status (effective in January-April 202l cohorts only) because these covariates were strong potential confounders. In the second step, a total of 38 binary, categorical, and continuous covariates were included in the propensity score model, including immunosuppressive medication use (binary), nursing home residence any time, and diagnosed CDC high-risk conditions:(11) coronary heart disease, cancer (excluding non-metastatic skin cancers), chronic kidney disease, congestive heart failure, pulmonary-associated conditions (including asthma, COPD, interstitial lung disease, and cystic fibrosis), dementia, diabetes, hypertension, liver disease, sickle cell/thalassemia, solid organ or blood stem cell transplant, stroke/cerebrovascular disorders, substance use disorder, anxiety disorder, bipolar disorder, major depression, PTSD, and schizophrenia.

Other categorical variables in the propensity score model included sex, race, ethnicity, rurality of the Veteran’s home ZIP code, state of residence, smoking status, and categorization of two comorbidity scores (CAN(12), Nosos(13)). Continuous covariates included age, body mass index (BMI), comorbidity score via Gagne index, distance from a Veteran’s home to nearest VA hospital, count of CDC high-risk conditions, count of mental health conditions, and four VA utilization measures (inpatient admissions, primary care visits, specialty care visits, mental health visits in the prior 2 years).

A caliper of 0.2 times the pooled estimate of the standard deviation of the logit of the propensity score was used to bound which uninfected patients could be matched to each infected patient.(14) To provide the survey team a sufficiently deep pool of matched controls to account for survey non-participation, the 25 matched uninfected patients closest in propensity score were retained for each infected patient. Infected patients with fewer than 25 matched uninfected patients had all their comparator patients selected as eligible matches. Matching was performed by the PSMATCH procedure from SAS/STAT 15.1 in SAS® 9.4M6 via the VA Informatics and Computing Infrastructure (VINCI) platform.

### Outcomes Comparisons to be Conducted

The EHR-based clinical outcomes that we intend to compare between matched cohorts are mortality, depression, suicide, onset of new clinical diagnoses, exacerbation of prevalent conditions, development of COVID-19 sequelae, health care use, and costs. The survey-based outcomes to be compared between matched cohorts include disability, financial toxicity, and health-related quality of life. For all analyses, we take a “per-protocol” approach such that uninfected patients who cross over to become infected will be censored at the time of infection. The study team discussed inclusion of negative control outcomes, but an outcome expected to be null between comparators could not be identified due to the ubiquitous effects of SARS-CoV-2 infection and the conditioning of negative control outcomes on health care utilization that might be differential between comparators.

## Results

From a sampling frame of 231,160 Veterans who had documentation of at least one SARS-CoV-2 infection between March 2020 and April 2021, and 9,291,822 Veterans without evidence of infection over the same time period, we excluded patients for missing comorbidity (CAN) score, no primary care use 24 months prior to index, missing or implausible height, weight, or age (Figure 1). We also excluded Veterans with missing ZIP codes or ZIP codes outside of Washington, D.C. or the 50 states, patients who were uninfected on the 1^st^ of each month but became infected later in the same month (for the uninfected group), or had a prior infection documented in Medicare. Lastly, we excluded 776 (0.4%) of 209,312 infected patients who did not have a suitable match, which generated final matched cohorts of 208,536 infected and 3,014,091 uninfected Veterans (comprising 5,173,400 total person-months of follow-up because of matching with replacement).

**Figure 1.**
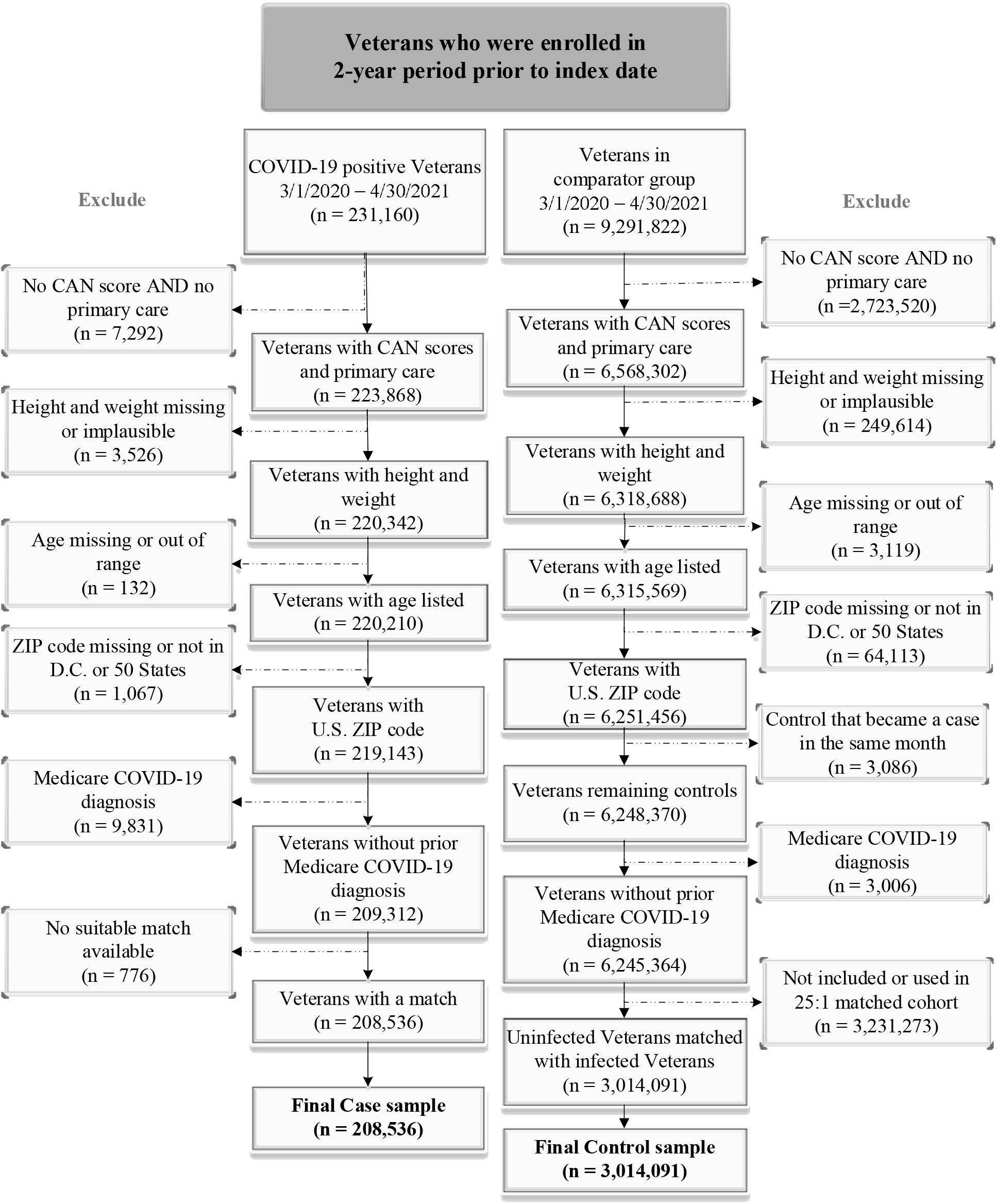
STROBE Figure of Cohort Derivation.

The matched cohorts were well-balanced on all covariates, based on standardized mean differences (SMD) <0.1 (Table 1). The cohorts included Veterans from all 50 states and Washington, D.C. Most Veterans in the matched cohorts were male (88.3%), non-Hispanic (87.1%), white (67.2%), and living in urban areas (71.5%), with a mean (standard deviation, SD) age of 60.6 (16.4), BMI of 31.3 (6.6), and mean (SD) straight-line distance to the closest VA medical center of 35.8 (35.2) miles. A minority were current smokers (12.6%), 39.3% had never smoked and 42.5% were former smokers. Comorbidity was assessed several ways, including Gagne score (mean=1.4, SD=2.2), count of CDC high-risk conditions (mean=2.3, SD=1.9) and count of 5 mental health conditions prevalent in Veterans (mean=0.9, SD=1.0). The most common diagnoses were hypertension (61.4%), diabetes (34.3%), major depression (32.2%), coronary heart disease (28.5%), PTSD (25.5%), anxiety (22.5%), and chronic kidney disease (22.5%). Approximately 10% of matched cohort members had been prescribed one or more immunosuppressive medications within 24 months before the index date. Of the 34.0% of the cohort with index dates between January-April 2021 when vaccinations became available, 70,985 Veterans (1.5% of the entire infected cohort) received at least one dose of a vaccine before their first positive COVID-19 test result.

**Table 1.**
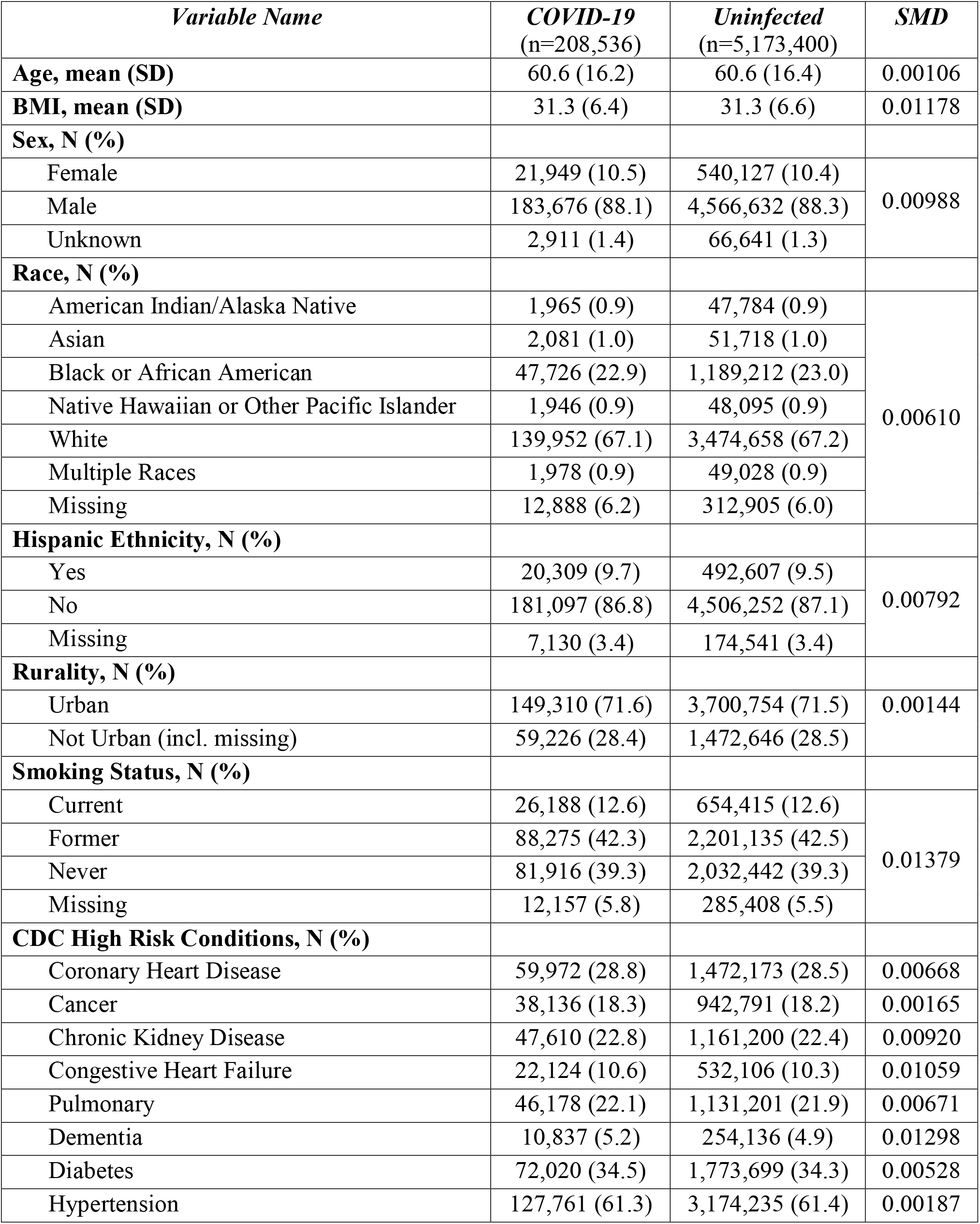
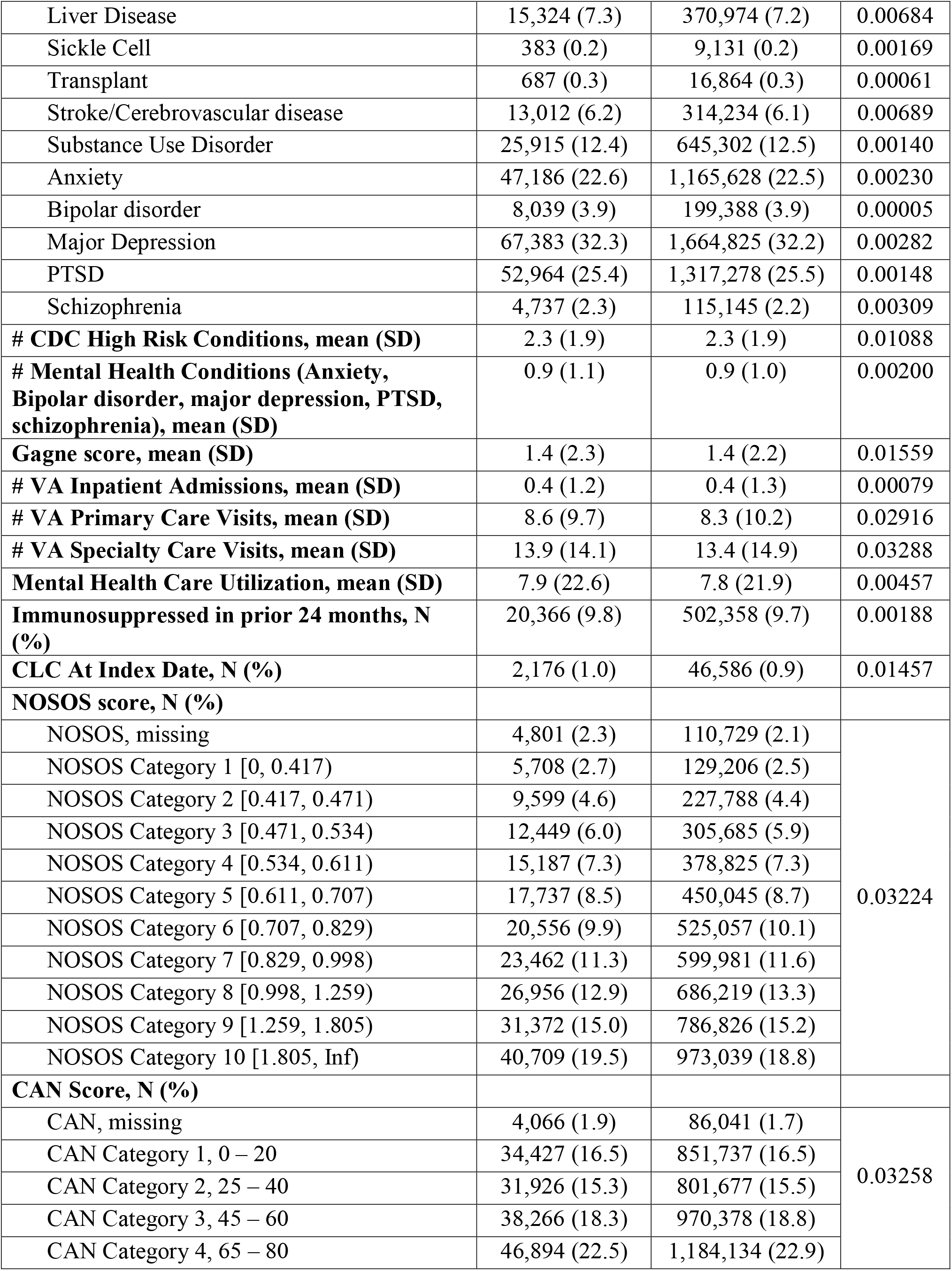
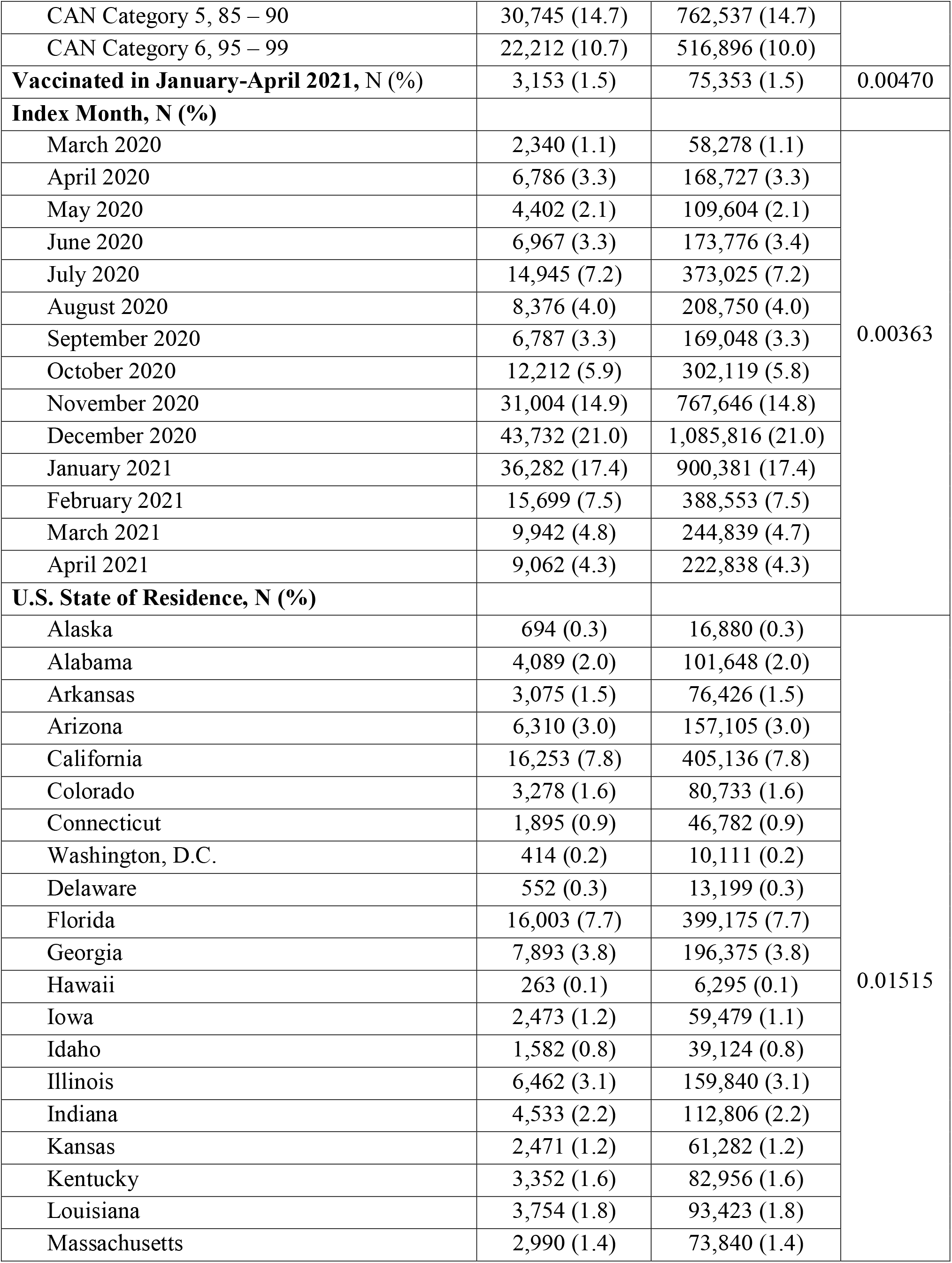
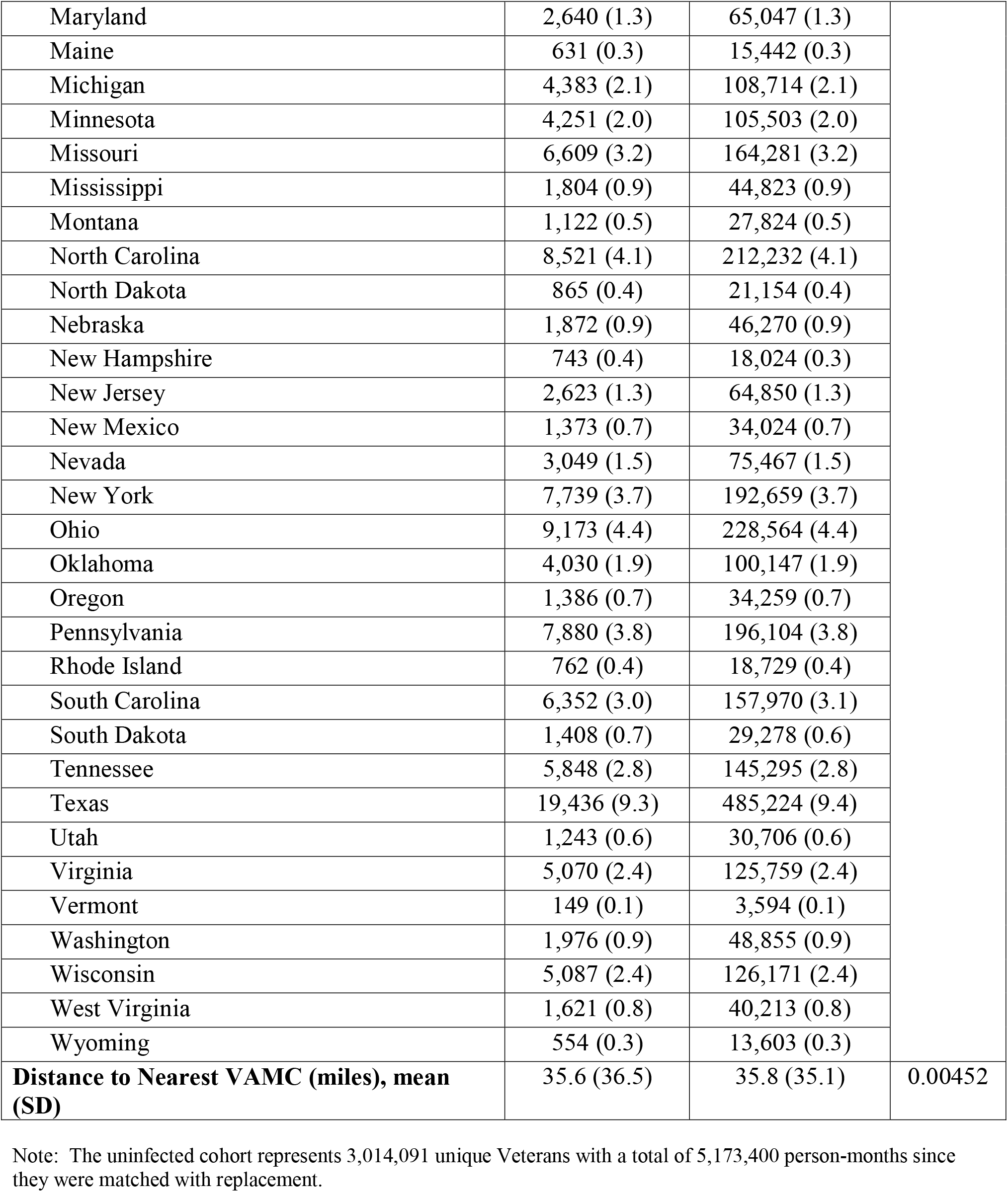
Descriptive Statistics of Matched Cohorts.

In the 24 months prior to the index date, cohort members had a mean (SD) of 8.3 (10.2) primary care visits, 13.4 (14.9) specialty care visits, and 7.8 (21.9) mental health visits in VA. Over one-half (53.3%) of infected patients were drawn from three of the 14 months in the observation period (November 2020, December 2020, and January 2021).

## Discussion

Despite the very large sample size available for this research with 231,160 infected and just over 9 million uninfected Veterans, a strategy of bias reduction based on coarsened exact matching on resulted in lack of an exact match for 53.7% of cases. The large sample loss reduced statistical power and generalizability since the exposure-disease associations may have differed between successfully matched and unmatched populations. The combined exact matching and propensity score approach, on the other hand, resulted in a much lower failure to match frequency at only 0.4%, with a high rate of success as assessed by the SMDs < 0.1 for all matching covariates. The work performed by the CORC to identify important covariates on which to and match cases and uninfected controls using propensity score methodology should facilitate the performance of causal research on long COVID-19 etiology in this population. Matched cohorts will be updated from May 2021 forward to be able to generate evidence on Veteran experience after April 2021.

As the nation’s largest national integrated publicly financed health system, the VA has the unique ability to track long-term outcomes among individuals infected with SARS-CoV-2 because it has a well-established comprehensive EHR that was developed around the mission of providing lifelong care for Veterans. In addition, Veterans are historically reliant on VA for care if they engage with the health system.

Our matching strategy defines the specific effect that will be estimated from our results. We considered historical controls of Veterans receiving care in the VA before the pandemic, but that would estimate the effect of individual SARS-CoV-2 infection combined with all the many other social and systematic disruptions that accompanied the pandemic. We also considered comparing Veterans hospitalized with SARS-CoV-2 infection to Veterans hospitalized with other conditions (e.g., influenza), which would be analogous to a randomized clinical trial with an active comparator. Such a comparator group asks whether COVID-19 hospitalization is worse than other sorts of hospitalizations. We reasoned that, for most Veterans, had they not developed COVID-19, they may not have been hospitalized with another condition that same month (although we did not exclude those hospitalized, so they do occur at whatever their natural frequency is in the comparator group).

We also did not wish to restrict to only hospitalized COVID-19 patients, as we took as a scientific question the relationship between initial severity of SARS-CoV-2 infection and subsequent outcomes—rather than presuming it by conditioning on initial severity. We also considered Veterans infected with other non-SARS-CoV-2 viruses. However, we noted the substantial body of evidence on sepsis and pneumonia—much of it viral in origin—that suggested such patients also have adverse long-term outcomes caused by non-SARS-CoV-2 viruses, including influenza. As such, we reasoned such comparators might underestimate the total individual effects survivors of COVID-19 would face and health systems would need to support. Each of these comparators may be of great interest to other research groups; they were not, however, our primary focus. Our goal was to generate matched cohorts to support cohort studies of EHR-derived outcomes and sample selection for qualitative interviews and patient surveys.

The retrospective cohort study described here is subject to several limitations. First, cohort matching results in sample loss that may reduce generalizability of results compared to weighting methods, although we were able to retain >99% of the infected patients in the sample after matching. Second, there is likely some contamination of the uninfected comparator group with Veterans with undiagnosed SARS-CoV-2 infection or who tested positive for SARS-CoV-2 with test results not available from private insurers, Medicare Advantage plans, Medicaid, or other community sources. Third, covariate specification for matching is based on our understanding of risk factors and confounders of SARS-CoV-2 infection as of spring 2022. Fourth, results may not generalize to Veterans who became infected after April 2021 or to non-Veterans. CORC will update matched cohorts from May 2021-March 2022, but that work is ongoing.

## Conclusions

Our understanding of the long-term outcomes of Veterans who were infected with SARS-CoV-2 will be gleaned from qualitative interviews, population-based surveys, and cohort studies of outcomes derived from EHRs. This study will explore all these approaches, all framed in the context of the matched cohorts generated from EHR data from the largest integrated health system in the U.S. Due to Veterans’ reliance on VA for care and eligibility for care once enrolled, we will be able to evaluate clinical and economic outcomes following their acute SARS-CoV-2 infection, as long-term outcomes two years after the onset of the pandemic are now being realized.

## Supporting information

SPIRIT checklist

## Data Availability

The datasets generated and/or analyzed during the current study are not publicly available due to U.S. Department of Veterans Affairs data restrictions prohibiting sharing. Contact the corresponding, Dr. Matthew Maciejewski, for data requests.

## ABBREVIATIONS

CAN: Care Assessment Need
CDC: Centers for Disease Control and Prevention
COPD: chronic obstructive pulmonary disease
CAD: coronary heart disease
HIV: human immunodeficiency virus
CKD: chronic kidney disease
CHF: congestive heart failure
SUD: substance use disorder
SMI: serious mental illness
PTSD: post-traumatic stress disorder
VA: Veterans Health Administration
CBOC: VA community-based outpatient clinic.

## DECLARATIONS

### Ethics approval and consent to participate

Initial and continuing reviews approved by were reviewed and approved by the Durham Veterans Affairs Institutional Review Board and Research and Development Committee. All methods were carried out in accordance with relevant guidelines and regulations. No informed consent was obtained because the project is secondary analysis of data. The Durham VA Institutional Review Board granted waivers for informed consent, HIPAA information, and HIPAA research on decedents.

### Consent for publication

Not applicable.

### Availability of data and materials

The datasets generated and/or analyzed during the current study are not publicly available due to Department of Veterans Affairs data restrictions prohibiting sharing. Contact the corresponding, Dr. Matthew Maciejewski, for data requests.

Matthew L. Maciejewski, Ph.D.

Department of Population Health Sciences

Duke University Medical Center

Durham, NC 27705

Phone: (919) 286-0411 ext. 5198

### Authors’ contributions

Conception: VAS, TSZB, PH, ESW, MN, JAP, KB, ASB, DH, EJB, GNI, TJI, CBB, AMO, MLM.

Design of the work: VAS, TSZB, PH, ESW, MN, JAP, ASB, DH, EJB, GNI, TJI, CBB, AMO, MLM.

Data acquisition: TSZB, KB, PG, AK, AF, AB, AH, TAS, GNI.

Analysis: VAS, TSZB, KB, MLM

Interpretation of data: VAS, TSZB, PH, MN, JAP, KB, ASB, DH, EJB, GNI, TJI, CBB, AMO, MLM

Drafted work: VAS, TSZB, MLM

Substantive revision: VAS, TSZB, PH, ESW, MN, JAP, KB, PG, AK, AF, AB, AH, TAS, ASB, DH, EJB, GNI, TJI, CBB, AMO, MLM

---

**Appendix 1.** Original and Final List of Covariates for Matching.

| *Original Covariate List* | *Final Matching Specification Used* |
| --- | --- |
| Sex | Sex |
| Use of immunosuppressive medication | Use of immunosuppressive medication |
| Age | Age |
| Race | Race |
| Ethnicity | Ethnicity |
| CAN comorbidity score | CAN comorbidity score (7 categories) |
| Nosos comorbidity score | Nosos comorbidity score (11 categories) |
| Gagne comorbidity score indicators | Gagne comorbidity score |
| Indicator of CDC high-risk conditions (Cancer other than non-metastatic skin cancer) | Indicator of CDC high-risk conditions (Cancer other than non-metastatic skin cancer) |
| Indicator of CDC high-risk conditions (COPD/asthma/interstitial lung disease/cystic fibrosis/pulmonary) | Indicator of CDC high-risk conditions (COPD/asthma/interstitial lung disease/cystic fibrosis/pulmonary) |
| Indicator of CDC high-risk conditions (hypertension) | Indicator of CDC high-risk conditions (hypertension) |
| Indicator of CDC high-risk conditions (diabetes) | Indicator of CDC high-risk conditions (diabetes) |
| Indicator of CDC high-risk conditions (Dementia) | Indicator of CDC high-risk conditions (Dementia) |
| Indicator of CDC high-risk conditions (CAD) | Indicator of CDC high-risk conditions (CAD) |
| Indicator of CDC high-risk conditions (HIV) |  |
| Indicator of CDC high-risk conditions (Liver disease) | Indicator of CDC high-risk conditions (Liver disease) |
| Indicator of CDC high-risk conditions (Pregnancy) | Indicator of CDC high-risk conditions (Pregnancy)<sup>1</sup> |
| Indicator of CDC high-risk conditions (Sickle cell/thalassemia) | Indicator of CDC high-risk conditions (Sickle cell/thalassemia) |
| Indicator of CDC high-risk conditions (Smoking) | Smoking status (4 categories) |
| Indicator of CDC high-risk conditions (Solid organ or blood stem cell transplant) | Indicator of CDC high-risk conditions (Solid organ or blood stem cell transplant) |
| Indicator of CDC high-risk conditions (Stroke or cerebrovascular disease) | Indicator of CDC high-risk conditions (Stroke or cerebrovascular disease) |
| Indicator of CDC high-risk conditions (CKD) | Indicator of CDC high-risk conditions (CKD) |
| Indicator of CDC high-risk conditions (CHF) | Indicator of CDC high-risk conditions (CHF) |
| Indicator of CDC high-risk conditions (SUD) | Indicator of CDC high-risk conditions (SUD) |
|  | Count of CDC high-risk conditions |
| SMI Diagnosis |  |
| Anxiety Diagnosis | Anxiety Diagnosis |
| PTSD Diagnosis | PTSD Diagnosis |
| Bipolar Diagnosis | Bipolar Diagnosis |
| Schizophrenia Diagnosis | Schizophrenia Diagnosis |
|  | Major depression Diagnosis |
|  | Count of mental health conditions |
| Body Mass Index (BMI) | Body Mass Index (BMI) |
| Community Living Center resident | Community Living Center resident |
| # VA telehealth visits |  |
| # VA in-person outpatient visits |  |
| # VA community care outpatient visits |  |
| # VA admissions | # admissions (VA + non-VA) |
|  | # VA primary care visits |
|  | # VA specialty care visits |
|  | # VA mental health care visits |
| State of residence | State of residence |
| Distance to nearest level 1a complexity VA facility | Distance to nearest VA medical center |
| Distance to nearest level 1b complexity VA facility |  |
| Distance to nearest level 1c complexity VA facility |  |
| Distance to nearest level 2 complexity VA facility |  |
| Distance to nearest level 3 complexity VA facility |  |
| Distance to nearest level CBOC |  |
| Rurality of residence | Rurality of residence (urban, rural) |
| Vaccinated for SARS-CoV-2 (January-April 2021) | Vaccinated for SARS-CoV-2 (January-April 2021) |
| Medicare enrolled (none, A only, A+B, A+B+D, MA) |  |

<sup>1</sup>Pregnancy indicator variable was constructed from ICD-10 diagnosis codes but was 0 for all patients in both arms of the study cohort.
Abbreviations: CAN: Care Assessment Need; CDC: Centers for Disease Control and Prevention; COPD: chronic obstructive pulmonary disease; CAD: coronary heart disease; HIV: human immunodeficiency virus; CKD: chronic kidney disease; CHF: congestive heart failure; SUD: substance use disorder; SMI: serious mental illness; PTSD: post-traumatic stress disorder; VA: Veterans Health Administration; CBOC: VA community-based outpatient clinic

